# Impact of dogs with deltamethrin-impregnated collars on prevalence of visceral leishmaniasis

**DOI:** 10.1101/19011577

**Authors:** Mondal Hasan Zahid, Christopher M. Kribs

**Affiliations:** University of Texas at Arlington, Department of Mathematics, Arlington, 76019, USA

## Abstract

Leishmaniasis is a vector borne zoonosis which is classified as a neglected tropical disease. Among the three most common forms of the disease, Visceral Leishmaniasis (VL) is the most threatening to human health, causing 20,000 to 30,000 deaths worldwide each year. Areas where VL is mostly endemic have unprotected dogs in community and houses. The presence of dogs usually increases VL risk for humans since dogs are the principal reservoir host for the parasite of the disease. Based on this fact, most earlier studies consider culling dogs as a control measure for the spread of VL. A more recent control measure has been the use of deltamethrin-impregnated dog collars (*DIDC*s) to protect both humans and dogs by putting *DIDC*s on dogs neck. The presence of dogs helps to grow the sandfly population faster by offering a more suitable blood-meal source. On the other hand, the presence of *DIDC*s on dogs helps to reduce sandfly population by the lethality of deltamethrin insecticide. This study brings an ecological perspective to this public health concern, aiming to understand the impact of an additional host (here, protected dogs) on disease risk to a primary host (here, humans). To answer this question, we compare two different settings: a community without dogs, and a community with dogs protected with *DIDC*. Our analysis shows the presence of protected dogs can reduce VL infection risk in humans. However, this disease risk reduction depends on dogs’ tolerance for sandfly bites.

## 1 Introduction

The Leishmaniases are a group of diseases caused by the protozoa parasite *Leishmania* [1, 3], which is transmitted by the bites of female sandflies [3, 4]. Over 20 Leishmania species known to be infective to humans are transmitted by the bite of infected female phlebotomine sandflies. Leishmaniasis is classified as a neglected tropical disease (NTD). It is found in parts of the tropics, subtropics, and southern Europe [3]. However, the disease mainly affects poor people in Africa, Asia and Latin America [1]. There are three main types of leishmaniasis among which visceral, often known as Kala-azar, is the most serious form of the disease [1]. Regarding visceral leishmaniasis, more than 90% of all cases occur in just the six countries of India, Bangladesh, Sudan, South Sudan, Brazil, and Ethiopia [13].

Out of 200 countries and territories reporting to the World Health Organization (WHO), 77 countries are endemic for visceral leishmaniasis in 2017. In 2016, over 90% of global VL cases were reported from seven countries: Brazil, Ethiopia, India, Kenya, Somalia, South Sudan and Sudan. As of October 2018, 50 VL-endemic countries have reported 2017 data to the WHO Global Leishmaniasis programme [1]. Annually, 700,000 to 1 million new cases and 20,000 to 30,000 deaths occur [2]. Currently, this is one of the major public health concerns. Even though human VL is spread by female sandflies, dogs are the main reservoir of the VL parasite. Thus, dogs presence in a community normally increases human VL incidence. In the last couple of decades, many clinical and mathematical studies have been conducted aiming to understand the dynamics of Zoonotic VL (ZVL). In these studies, researchers tried to find ways of controlling VL prevalence. Different strategies for controlling the incidence of VL have been considered, most notably culling dogs and putting insecticide-impregnated dog collars on dogs.

In 2002, Gavgani et al. clinically studied two possible strategies– early diagnosis and treatment, and use of deltamethrin-impregnated dog collars (*DIDC*). The presence of deltamethrin insecticide on dog collars helps to kill a portion of sandlies, and eventually reduces the disease transmission. Outcomes of the study showed that use of *DIDC* is helpful in reducing human infection, and this strategy can replace the controversial dog culling program [8]. In addition to clinical studies, several mathematical models have also been used to study transmission and control strategies for ZVL. In 2013, Ribas et al. proposed and analyzed a deterministic mathematical model in order to compare different control strategies. They showed that using *DIDC* is better at reducing human infections[5].

In 2016, Zhao et al. studied ZVL transmission using an SEIR deterministic model [9]. In their model they incorporated hospitalization of infected humans, migration for the sandfly population, and infection-related death for sandflies. Also, they assumed that the contact rates between sandflies and hosts (dogs and humans) are independent of vector population, biting exposed dogs cannot infect sandflies, and immunity for both of the hosts, humans and dogs, is permanent. Later, they compared three different control strategies − vaccination of dogs, use of insecticide at vector breeding sites, and personal protection. Their analysis found controlling the sandfly population to be the most effective control measure.

The next year, in 2017, Shimozako et al. used the SEIR deterministic model also to study ZVL disease dynamics. They included a delay term instead of using latent compartment for the sandfly population [6]. In this study, they estimated the basic reproduction number *R*_0_ and analyzed the stability and sensitivity of the system, and finally made some recommendation regarding control strategies. Unlike Zhao’s work, they assumed that sandflies can be infected from biting exposed dogs, and immunity gained by both of the host populations is temporary. Interestingly, they assumed that exposed dogs and humans become susceptible to VL when recovery precedes the appearance of symptoms. Their work also assumed that the rate at which vectors bite hosts is independent of host density. Outcomes of their study showed that control strategy for ZVL should be focused on sandflies and infected dogs. However, considering the ethical concerns regarding culling dogs they recommend to prioritize the control of sandfly population.

All the research related to ZVL has mainly addressed public health concerns where researchers study different control strategies. In these studies, we always find the presence of human and dog populations as hosts for sandflies, the vector of the disease. This host richness (host diversity) leads us to think about the dilution effect (reduction in disease risk resulting from species diversity). The effect of the presence of an additional host is not straightforward. It can increase, or decrease disease risk depending on varieties of factors. In 2010, Johnson and Thieltges showed that host diversity helps in reducing human infections depending on the relative abundance of additional host(s) relative to the focal host [10]. In 2014, Miller and Huppert (2014) proved that species diversity in host population can amplify or can dilute disease prevalence depending on vectors’ preference of host [11]. Recently in 2019, Zahid and Kribs established that the presence of an additional host in domestic settings can help to reduce disease prevalence in humans if the distance between two host populations remains within a certain range [12]. These works challenge the established idea that biodiversity always helps to reduce disease risk. It is always interesting to observe how the presence of other hosts, in addition to humans, influences the dynamics of vector-borne diseases, impacts disease risk and human health.

This study shifts the research question from a public health viewpoint to an ecological one. As dogs are the main reservoir for the parasite, the usual presence of dogs in a domestic, or in a community setting makes VL transmission faster ensuring more suitable blood-meal source for its vector sandflies. Hence, this paper aims to identify the impact of the presence of protected dogs (protected by putting deltamethrin-impregnated dog collars) as an additional host on the prevalence of VL in humans.

The presence of protected dogs in a community has two contradictory effects. It ensures a better blood-meal source for vector population since biting a dog is much easier for sandflies than biting a human. Thus, dog presence in a community helps sandfly population to grow faster. Moreover, dog presence increases the proportion of infected sandflies (since dogs are the main reservoir of the parasite) which eventually increases human infection risk. On the contrary, use of *DIDC*s on dogs as topical insecticide reduces the sandfly population by the lethality effect, which can result in fewer cases of VL. The net result of these two contrary effects may reduce or enhance the risk of human prevalence. The goal of this study is to understand and examine this net effect, and eventually to understand if the presence of protected dogs has any dilution effect on human risk of VL infections. To answer this question, here we consider two different settings: a setting with protected dogs, and a setting with no dogs. To analyze and understand these two different settings, in this study we use an SEIRS deterministic model.

## 2 Model development

The SEIRS model we use here to understand the dynamics of VL incorporates three different populations: two hosts − dogs and humans, and the vector − sandflies. We neither consider hospitalization of sick humans, nor any migration for the vectors. Based on results of an early clinical study [14], we include disease transmission from symptomatic infected humans to vectors. This inclusion makes this model different from the models formulated by Zhao et al. [9], and by Shimozako et al. [6].

Like the models proposed by [5, 6], we also assume hosts’ immunity temporary. Similar to their model, we assume both of the hosts may acquire natural immunity directly from the exposed state. In contrast to [5, 6], however, we assume that exposed hosts cannot become susceptible without having any immunity. In our model, we have 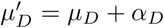 and 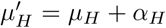 where *α*_*D*_ and *α*_*H*_ represent VL-induced death rates for dogs and humans respectively. The presence of deltamethrin on the collars causes additional deaths at the rate *α*_*S*_ (migration of sandflies due to presence of *DIDC*s, if any, can be included in *α*_*S*_) for vectors. So, the vectors are leaving at a rate 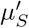 where 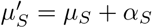.

The most important factor which makes our model distinct from others’ models is the encounter (biting) rate between hosts and vectors, which incorporates the notion of host irritability. The maximum number of bites per unit time a dog can tolerate is not the same as the maximum number of bites per unit time a human can tolerate. This issue of host-density dependent encounter rate is addressed by Blayneh et al. in 2010 [16] while modeling dynamics of West Nile Virus. However, the contact rate they used is independent of host population size. But, the sandfly biting rate is limited both by the sandfly’s preferred feeding rate and by host irritability (unlike mosquitos, which are limited primarily by availability of breeding sites). So, we consider the encounter rate between sandflies and dogs 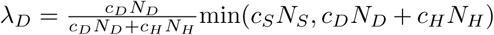, and the encounter rate between sandflies and humans 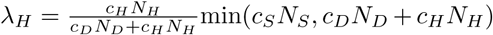 where *c*_*D*_ and *c*_*H*_ represent the number of bites a dog, and a human can tolerate per unit time, *c*_*S*_ represents the number of bites a single sandfly desires to make per unit time, and *N* ‘s represent population sizes. This inclusion of the host population dependent biting rate makes our model distinct from other earlier proposed models. However, not all the bites (encounters) can transmit the disease and so we multiply the total number of encounters by *b*_*D*_ (or *b*_*H*_ or *b*_*SH*_ or *b*_*SD*_) (Table 2) which represents the proportion of bites (between 0 to 1) that result new infections to dogs (or to humans or to sandflies). Finally, we have our model which is shown in Figure 1 and described by system (1). All the model variables are summarized in Table 1.

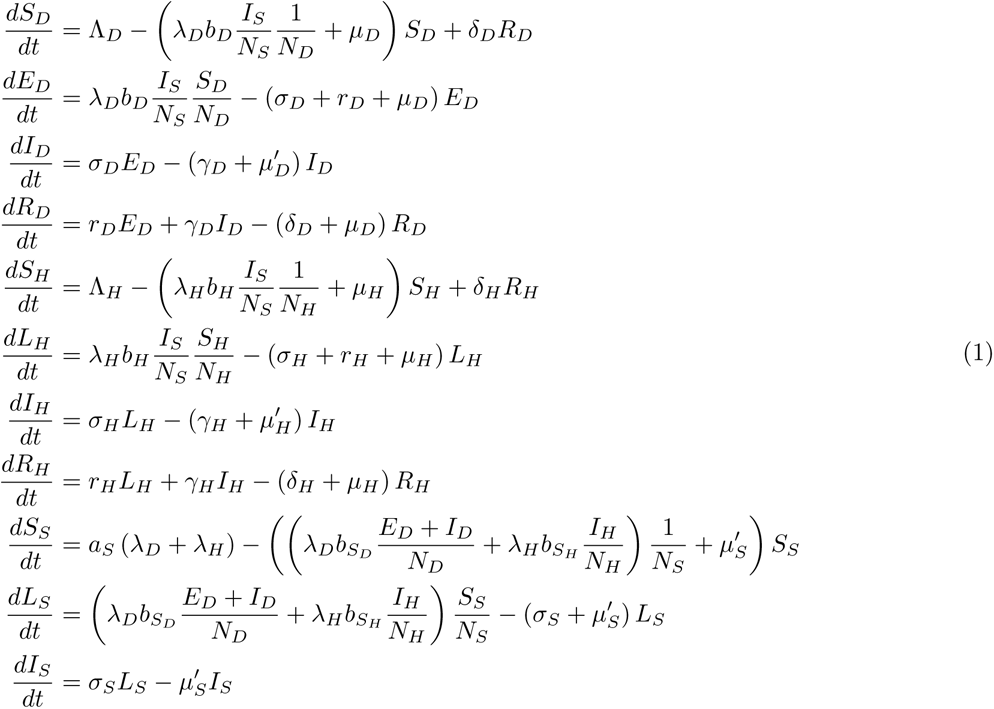

where

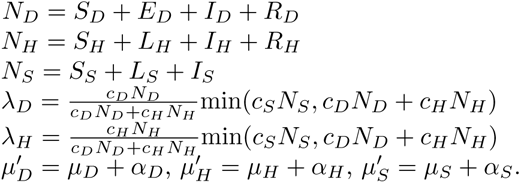

**Table 1:** Model variables with definition

| Variable | Definition |
| --- | --- |
| $S_D, S_H$ | Susceptible dogs, Susceptible humans |
| $L_H, L_S$ | Latent humans, Latent sandflies |
| $E_D$ | Exposed dogs |
| $I_D, I_H$ | Infected humans |
| $S_S$ | Susceptible sandflies |
| $I_S$ | Infected vectors |
| $R_D, R_H$ | Recovered (Temporary) Dogs, Recovered (Temporary) humans |

**Table 2:** Summary of model parameters

| Par. | Definition | Value | Units | Reference |
| --- | --- | --- | --- | --- |
| $\Lambda_D$ | recruitment rate for dogs (by birth) | 8.39 | dogs/day | This study |
| $\Lambda_H$ | recruitment rate for humans (by birth) | 1.42 | humans/day | This study |
| $a_S$ | birth rate for sandflies | 10 | sandflies/bite | This study |
| $b_D$ | infection to dogs from sandflies' bite | 0.01 | infected dog/bite | [17] |
| $b_H$ | infection to humans from sandflies' bite | 0.5 | infected human/bite | [17] |
| $b_{SD}$ | infection to sandflies from biting dogs | 0.3025 | infected sandfly/bite | This study |
| $b_{SH}$ | infection to sandflies from biting humans | 0.148 | infected sandfly/bite | This study |
| $c_D$ | inverse of dogs' irritability | 45 | bites/dog-day | This study |
| $c_H$ | inverse of humans' irritability | 15 | bites/human-day | This study |
| $c_S$ | bites a single sandfly disere | 0.13 | bite/sandfly-day | This study |
| $\sigma_D$ | incubation rate | $1.11 \times 10^{-2}$ | 1/day | This study |
| $\sigma_H$ | incubation rate | $6.85 \times 10^{-3}$ | 1/day | [31] |
| $\sigma_S$ | incubation rate | 0.117 | 1/day | This study |
| $\gamma_D$ | recovery rate (dogs) | $9.04 \times 10^{-4}$ | 1/day | [26] |
| $\gamma_H$ | recovery rate (humans) | $2.5 \times 10^{-3}$ | 1/day | [6] |
| $\delta_D$ | Inverse of temporary recovery period (dogs) | $2.74 \times 10^{-3}$ | 1/day | [28] |
| $\delta_H$ | Inverse of temporary recovery period (humans) | $5.48 \times 10^{-4}$ | 1/day | [28] |
| $r_D$ | spontaneous recovery rate | $1.1 \times 10^{-2}$ | 1/day | [27] |
| $r_H$ | spontaneous recovery rate | $8.22 \times 10^{-3}$ | 1/day | [26] |
| $\mu_D$ | natural death rate | $2.30 \times 10^{-4}$ | 1/day | This study |
| $\mu_H$ | natural death rate | $3.6 \times 10^{-5}$ | 1/day | This study |
| $\mu_S$ | natural death rate | 0.0714 | 1/day | This study |
| $\alpha_D$ | disease induced death rate for dogs | $1.14 \times 10^{-3}$ | 1/day | This study |
| $\alpha_H$ | disease induced death rate for humans | $8.26 \times 10^{-7}$ | 1/day | This study |
| $\alpha_S$ | *DIDC* induced death rate for sandflies | 0.1995 | 1/day | This study |

**Figure 1:**
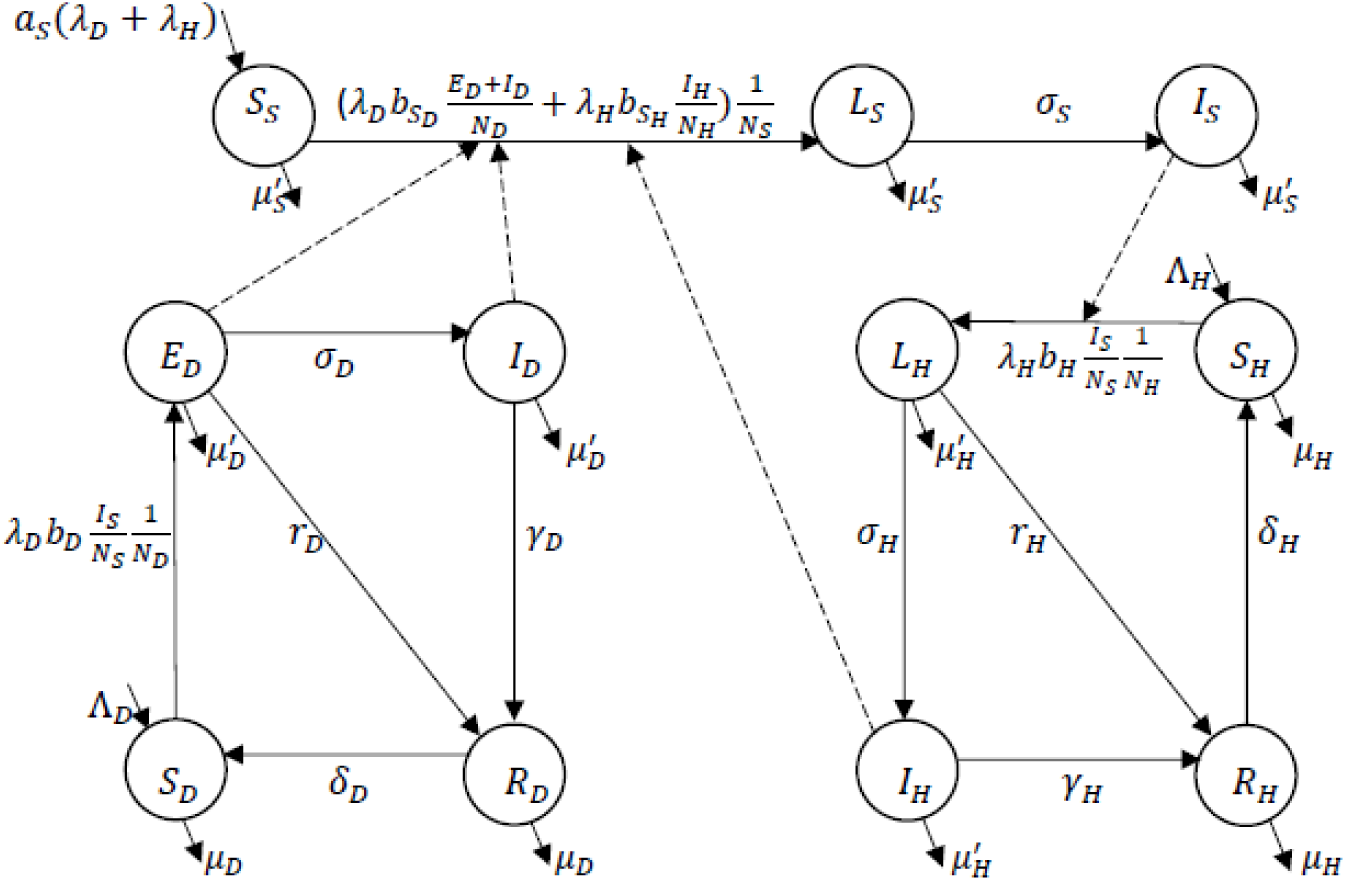
Population flow among the compartments

## 3 Parameter Estimation

In 2017 the World Bank listed 207,833,831 as total population and 13.918/year as the birth rate (crude) per 1000 people for Brazil [19]. We use the total number of municipalities in Brazil, 5570 [20], to estimate the average population (37,313) in a single municipality of Brazil. Then using this average population and the birth rate we estimate the recruitment rate for humans to get Λ_*H*_ =1.42 humans/day. To estimate the recruitment rate in the dog population, we use the results of a study performed in 2005-2008 in Vargem Grande, a neighborhood of the municipality of São Paulo, Brazil, with a population of 16,946 [21]. The study estimated 1337 and 1445 new dogs in 2006 and 2008 respectively, which gives a mean increase of 1391 dogs/year. Then we apply the ratio 2.20, of the population per municipality (estimated above) to the population of the study area of [21], which estimates an increment of 3060 dogs/year in a municipality of average population. And, we get 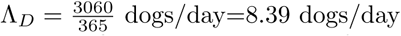.

Female sandflies take 3 to 5 days after their emergence to take a blood-meal, and it takes 7.67 days (mean of 6 days, 8 days, and 9 days) from blood-meal to oviposition (laying of eggs) [15]. They usually take only one blood-meal until they lay eggs, and begin feeding again after oviposition [34]. Therefore, each vector has a single bite in every 7.67 days, and thus we have 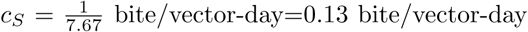. Also, we have 10 (mean of 13, 8 and, 9) new female sandflies per egg batch [15] which gives us *a*_*S*_=10 sandflies/bite.

A study in 2010 found 12 sandflies infected when 81 sandflies were fed on people with active VL infection [14], and this gives us 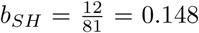 infected sandfly/bite. Another study in 2013 showed that 35.8% of sandflies that fed on asymptomatic dogs, and 24.7% of sandflies that fed on symptomatic dogs got the infection [18]. We take the mean (30.25%) of these two estimations to get *b*_*SD*_ =0.3025 sandfly/bite. However, for *b*_*H*_, *b*_*D*_ we take the estimations from [17] as 0.5 infected human/bite, and 0.01 infected dog/bite respectively.

In 2017, life expectancy at birth in Brazil was 76 years [19], so we take its reciprocal to estimate the natural death rate for people in Brazil which gives *µ*_*H*_ = 3.6 × 10^−5^/day. In Brazil, there are four common dog breeds, and their mean lifespan is 11.875 years [22]. We take the reciprocal of the life span as death rate, and we get *µ*_*D*_ = 2.30 × 10^−4^/day. To estimate the natural death rate of sandflies we take the reciprocal of their mean lifespan which is 2 weeks [23], and get *µ*_*S*_ =0.0174/day.

A study of ZVL cases in Bihar, India, in 2013 showed that 154 patients died among a total of 3,641 patients [24], and their average life span was 66.73 years. However, the life expectancy at birth in Bihar at that time period was 68.1 years [25]. Therefore, we take the difference of the reciprocals of these two life span and estimated lethality of ZVL as 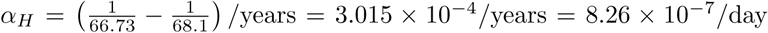. Earlier studies show that average life-span of infected dogs is two years [17, 35] which gives us 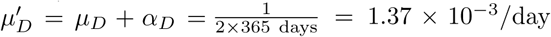. This value, and already estimated value *µ*_*D*_ = 2.30 × 10^−4^/day give us 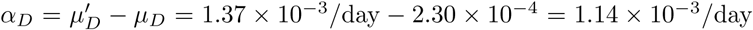. To estimate the *DIDC* induced death rate (*α*_*S*_) for sandflies, we study [29] where sandflies were exposed to dogs protected (with *DIDC*). From each individual experiment, we take the number of exposed sandflies, and the number of flies that died in 20 hours duration from their exposure, and pooled data from all trials to get 5,766 and 1245 as the totals of exposed sandflies, and dead sandflies respectively. However, our estimation of sandflies’ natural death rate (*µ*_*s*_=0.0714/day) estimates 333 natural deaths ^1^ of sandflies in a span of 20 hours. The remaining deaths of 1245-333=912 of sandflies not attributable to natural mortality. These estimations, and sandflies’ natural death rate 0.0714/day give us the estimation 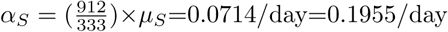.

A laboratory study in 2011 observed sandflies’ incubation rate as 7-10 days [30]. So, here we take the reciprocal of the mean of 7 and 10 days to estimate the latent period as 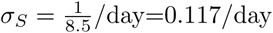. Another work in the same year estimated that individuals without symptomatic VL need on average about 146 days to develop LST-positivity after a PCR-positive finding [31], and this leads us to estimate 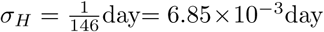. A review paper published in 2002 mentioned the incubation period for dogs as 2-4 months [32]. We took reciprocal of the mean of this range of 2-4 months (3 months=90 days), and estimate 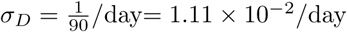.

All other parameter values are taken directly from earlier research studies.

## 4 Analysis

We begin with the general model where we incorporate three different populations – sandflies (the vector), and two host populations– dogs (protected with *DIDC*) and humans. Later, we consider a special case without dogs in the setting. Finally, we compare these two cases to understand the impact of the presence of protected dogs on the prevalence of VL infections in humans. The study actually aims to understand the effects of host diversity on disease transmission, and eventually on human health.

In the general model, the total number of desired bites for sandflies and the total number of bites that hosts can tolerate together may not be equal. Thus, in our analysis we consider two cases in terms of total number of possible encounters. In the first case, we assume the maximum total number of possible vector bites is less than the total possible number of bites that both hosts (humans and protected dogs) can tolerate together, that is when *c*_*S*_*N*_*S*_ < *c*_*D*_*N*_*D*_ +*c*_*H*_*N*_*H*_. The other scenario takes place when *c*_*S*_*N*_*S*_ > *c*_*D*_*N*_*D*_ +*c*_*H*_*N*_*H*_. For the initial case our calculation estimates the total vector population as 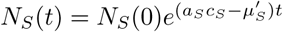 (recall 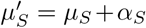) which shows that the vector population decreases with time and eventually dies out, if 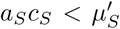. However, *N*_*S*_ increases with time under the condition 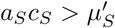. If the vector population continues to grow then the base condition of the first case, that is *c*_*S*_*N*_*S*_ < *c*_*D*_*N*_*D*_ + *c*_*H*_*N*_*H*_, cannot be true after a certain time. Eventually, the relation between the maximum possible number of vector bites and the maximum host bite tolerance (inverse of hosts’ irritability) turns into *c*_*S*_*N*_*S*_ > *c*_*D*_*N*_*D*_ + *c*_*H*_*N*_*H*_, which is the second case. These analyses give us two scenarios: either the vector population dies out, or the only possible case is *c*_*S*_*N*_*S*_ > *c*_*D*_*N*_*D*_ + *c*_*H*_*N*_*H*_ (second case). Since we are interested to understand the disease dynamics, from here our study will consider only the case of *c*_*S*_*N*_*S*_ > *c*_*D*_*N*_*D*_ + *c*_*H*_*N*_*H*_.

To find equilibrium points for our model we set all the equations of our system equal to zero and solve them. After performing some basic arithmeticc we get the disease free equilibrium (*DFE*) as 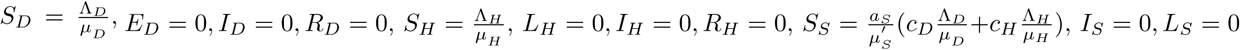. Then we use the next generation method [33] to get the basic reproduction number (*BRN*),

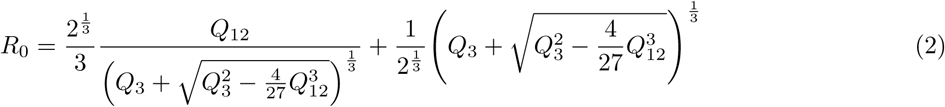

where

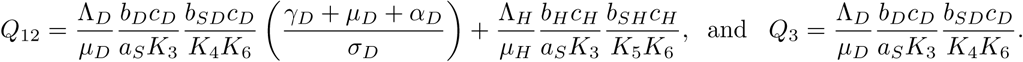

Here *Q*_12_ accounts for transmission between infected vectors and infected hosts, while *Q*_3_ accounts for transmission between infected vectors and exposed dogs. The terms are not simply added to form *R*_0_ because the cycles overlap as exposed dogs become infected dogs later. However, when 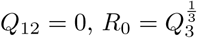, and when 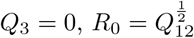. In general 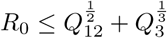.

Later, we obtain the following quadratic equation (in *I*_*H*_) for endemic equilibrium:

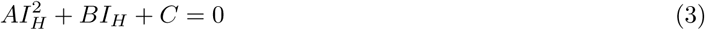

where

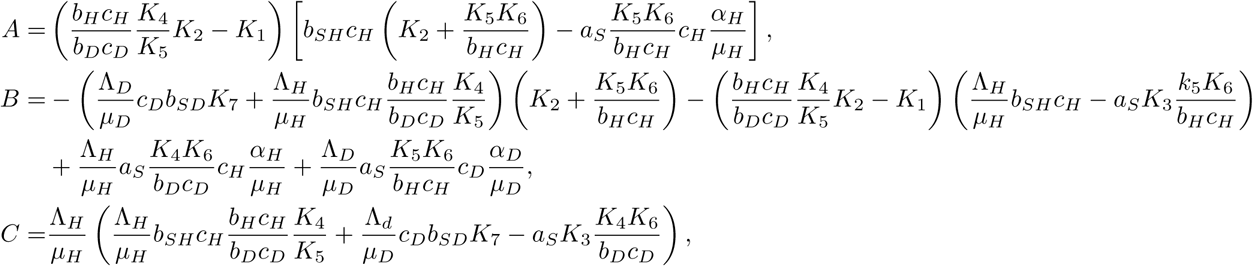

with

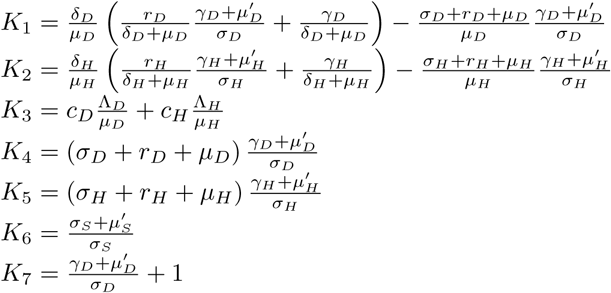

To understand the behavior of disease dynamics, the threshold value *BRN* (*R*_0_) and endemic equilibrium (*EE*) need to be understood, and interpreted properly. Our analysis shows *R*_0_ > 1 if and only if *C* > 0. For endemic equilibrium, we cannot establish any specific condition to ensure the positivity of *EE* since its analytic expression, obtained from equation (3), is very complex (recall expressions of coeffficients *A, B* and *C*). Consequently, we perform numerical explorations to understand the behavior of our dynamical system, and finally conclude that the model has a unique endemic equilibrium whenever *R*_0_ > 1. It does not appear possible for *A, B, C* all to be positive together. To check the stability of our *EE*, we evaluate the Jacobian matrix of our dynamical system, and perform numerical explorations. Our simulations show that the endemic equilibrium is unconditionally stable.

Now, we consider the case of having no dogs in the scene, a special case (setting all variables and parameters related to dogs to zero) of our original model. Our analysis for this special case finds the *DFE* as 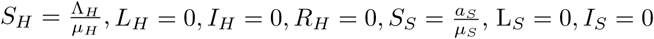, and *BRN* as 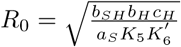 where 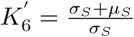 (derived from *K*_6_ by setting *α*_*S*_ = 0). For the endemic equilibrium, we get the expression for the infected human population as

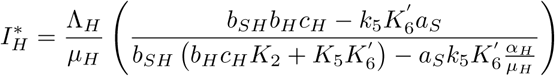

which can be expressed in terms of *R*_0_ as

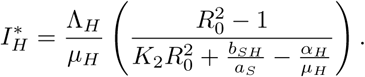

Based on our parameter estimation, we calculate *R*_0_ and 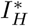 for all three possible cases: community with dogs protected with *DIDC*, community without dogs, and community with dogs having no protective measure. We estimate 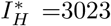, 3329, and 4677 out of 39,445, respectively, for these three cases. These estimates indicate that the presence of DIDC-protected dogs is better than having no dogs, which in turn is better than having unprotected dogs (in terms of human infections of leishmaniasis). The *R*_0_ values for these same cases are 1.47, 3.51, and 2.05 respectively. These results provide fewer human infections with a higher *R*_0_ value for the no dog case compared to the case of unprotected dogs. Humans are spreading infections faster in the case of no dog; however, the contribution of the dog population to new infections is missing in this case. Thus, the case of no dog in the community produces fewer infections even though the *R*_0_ value is higher compared to the case of unprotected dogs. The expression for *R*_0_ in (2) helps us to understand this apparently unusual result better. One of *Q*_12_’s two terms, and all of *Q*_3_, have to do with dogs. Removing them will reduce *R*_0_, especially since sandfly biting rate is asymptotically host-dependent. Figure 2 shows how the order of *R*_0_ values changes for a certain parameter (*c*_*H*_) range, to match the 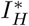 ordering.

**Figure 2:**
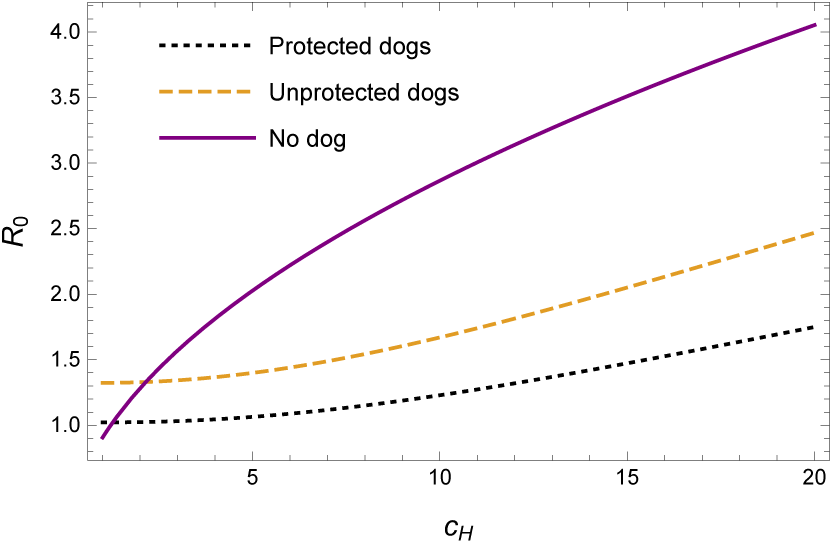
*R*_0_ values change their order while a parameter (*c*_*H*_) varies

A local sensitivity analysis of the endemic prevalence of leishmaniasis 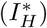, for the case of protected (with *DIDC*) dogs’ presence, was performed for all of our model parameters (Figure 3). Values for the 6 parameters with normalized sensitivity indices greater than 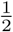 are well known, and of the parameters with normalized sensitivity indices greater than 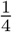, the two most of interest to this study are *c*_*D*_ and *α*_*S*_.

**Figure 3:**
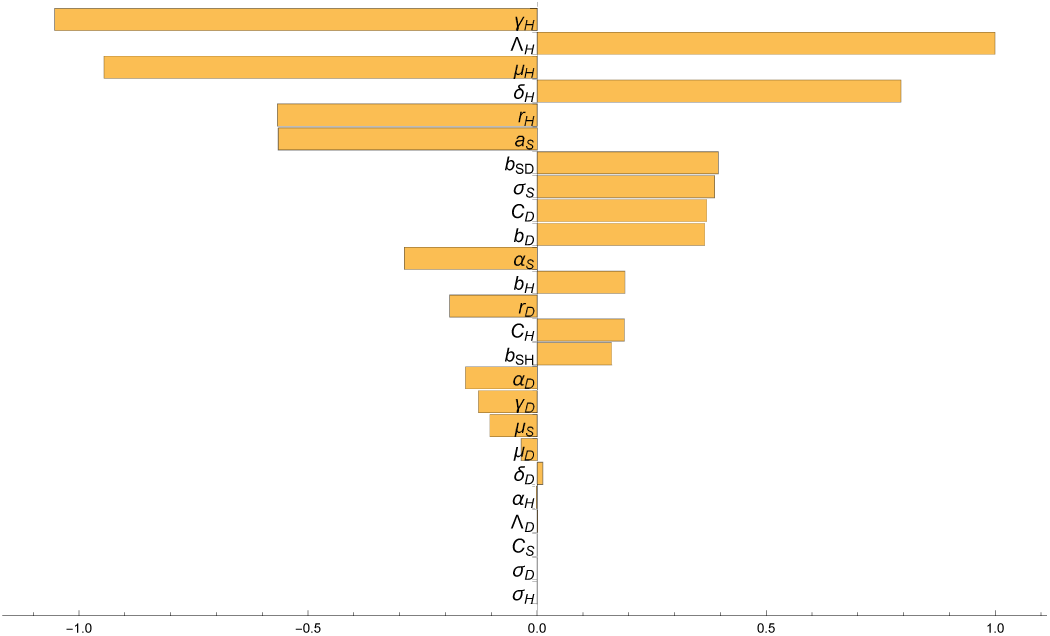
Local sensitivity analysis of 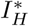 for all model parameters

Now we vary the values of *c*_*D*_ and *α*_*S*_ together to observe the contribution of dogs’ irritability and the efficacy of *DIDC* simultaneously in reducing human infections. Our analysis shows that the number of human infections increases with dogs’ tolerance for bites (*c*_*D*_), because this allows vectors’ easy access to bite dogs which helps the sandfly population to grow. It also increases the proportion of infected sandflies, because the probability of infection to sandflies from biting dogs is higher than the probability of new infection from biting humans. The number of human infections increases also for extremely low dog tolerance, because it reduces the lethality of protected collars by reducing the number of interactions between sandflies and dogs significantly. Also, humans get almost all the bites here which eventually increases human infection risk. Hence, the number of human cases of VL infections is not always proportional to dogs’ tolerance to sandfly bites. Figure 4 clearly explains the above discussion regarding the effect of dogs’ tolerance for sandfly bites. Our analysis also shows that human infections decrease with the population size of protected dogs. However, based on our parameter estimation, we find 58% of the dog population needs to be protected with *DIDC* to ensure the effectiveness of the presence of protected dogs in reducing human risk of VL infections.

**Figure 4:**
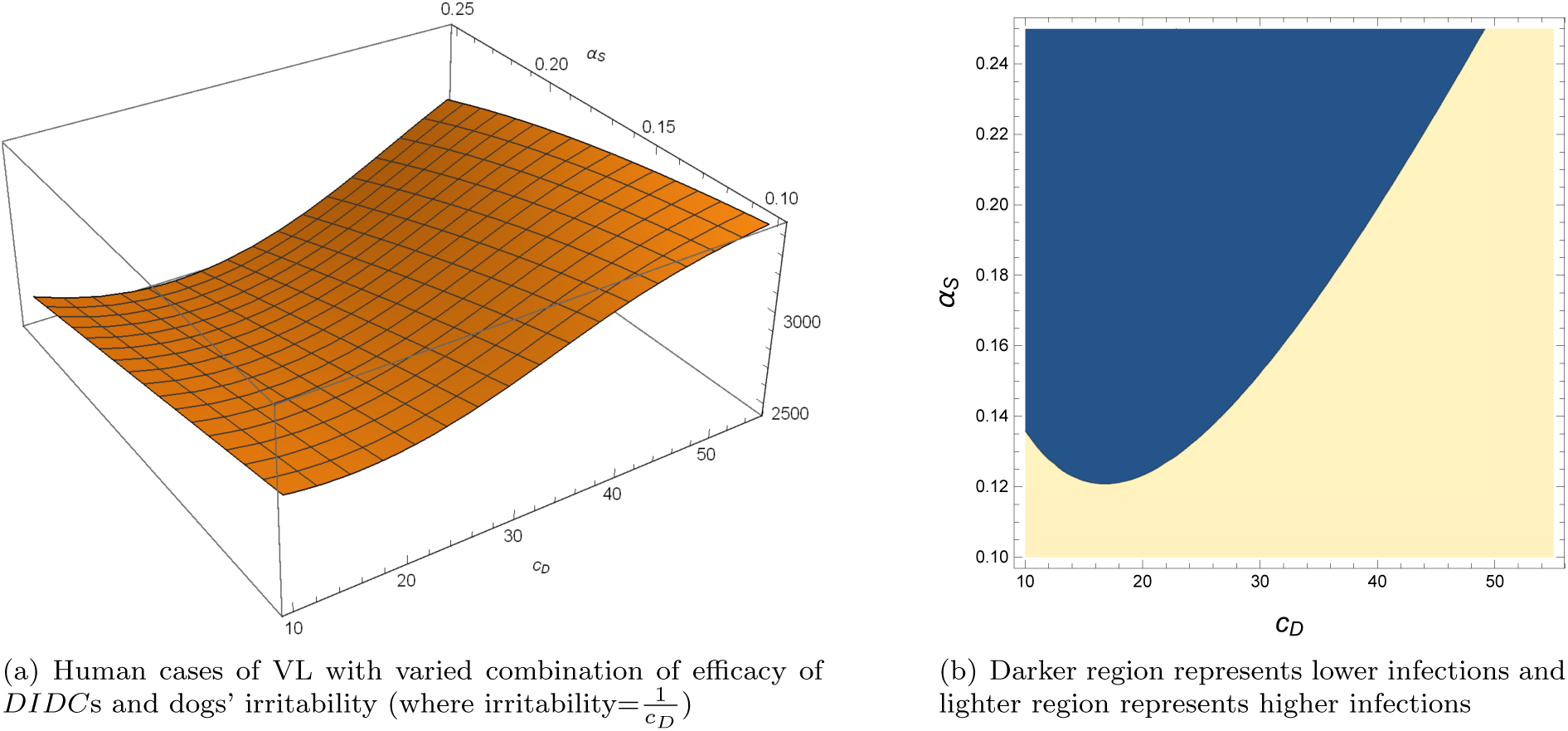
Effect of dogs’ irritability, and *DIDC* efficacy on human infections of VL

## 5 Discussion

The model used in this study gives a new insight to the study of visceral leishmaniasis transmission among human, dog, and sandfly populations. This happens as this study considers humans as a competent host (based on earlier research [14]), in contrast to other studies’ assumption of treating humans as a dead-end host (assuming dogs as the only source of vector infections). The host-population-dependent biting rate for sandflies highlights the impact of host biodiversity more than other models used in the earlier studies of leishmanisis. Our results show the presence of dogs with *DIDC* (as topical insecticide) in a community produces fewer human infections compared to infections in the same community without dogs. It is also confirmed that a community without dogs is better in terms of VL infections in humans than a community containing unprotected dogs.

Zhao et al. (2016) [9] develop and analyze an SEIR model assuming hosts’ recovery from infections permanent. In our model, we consider recovery for hosts as temporary which leads us to use an SEIRS model. They incorporate sandfly migration in the model which we do not. Their analysis finds the condition *R*_0_ < 1 insufficient for complete control of the disease since they observe the existence of backward bifurcation under the condition *R*_*c*_ < *R*_0_ < 1. In our study, we find only one endemic equilibrium which precludes any possibility of the existence of backward bifurcation. Identifying which feature of their model is responsible for the backward bifurcation is prevented by the fact that their study did not provide the definition of *k*_3_ which is present in the definition of *R*_*c*_. Even though our study does not focus on optimal control policy like Zhao et al., we study the impact of presence of protected (with *DIDC*) dogs, and find this presence helpful in producing fewer human infections.

Shimozako et al. (2017) [6] assumed recovery for both hosts from VL infections is temporary, so they used an SEIRS model in studying ZVL transmission. We also use a modified SEIRS model even though we exclude their assumption that exposed hosts (dogs and humans both) can become susceptible without developing any immunity. We also do not adopt their assumption that latent, and clinically ill dogs have different probabilities of infecting sandflies. They find VL transmission completely dependent on the dog and sandfly populations. However, our study shows each of the three populations has contributions in the dynamics of VL transmission. Shimozako et al. (2017) suggests that control of VL transmission should be based on the sandfly population. Our analysis agrees with this suggestion, showing that presence of dogs with *DIDC* protection (which reduces sandfly population) reduces human cases of visceral leishmanisis.

Our study also draws on an ecological perspective to inform public health policy. In the literature review, we mentioned ecological studies [10, 11, 12] in which the presence of additional hosts (known as host richness) may help in reducing the human infection risk depending on some factors, such as the abundance of additional host(s) relative to the focal host, vectors’ preference of host for feeding, and distance between the primary host and additional host. This study also found the presence of an additional host (dogs protected with *DIDC*) helpful in reducing human risk of VL infections. However, this reduction is independent of all three of the factors which are identified in [10, 11, 12]. We find that dogs’ presence in a community does not produce fewer human infections if dogs’ irritability is very high, or extremely low even after *DIDC*s are ensured on them. This ecological change helps to protect human health only if dogs’ irritability ranges somewhere in the intermediate level.

Our proposed model has a few limitations in its development. In our study, we assume all dogs in a community are protected by deltamethrin-impregnated collars which may not be possible in reality. Also, we do not incorporate sandfly migration in our model. However, this migration rate can be included in our *DIDC*-induced sandfly death rate (*α*_*S*_). Inclusion of this migration may have some impact on our numerical results, and also on the range of dog irritability values which are helpful. However, our qualitative results will remain the same. Addressing this limitations could produce better insights into visceral leishmaniasis dynamics. Our proposed model offers a better base than other models for studying control strategies for ZVL, and VL transmission since we incorporate some real, and very important issues, like human infectivity and the role of host irritability.

## Data Availability

N/A

## 6 Declaration of competing interest

The authors declare no conflict of interest.

## Footnotes

1 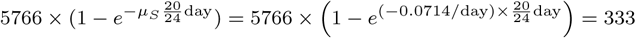

## Notes

### Competing Interest Statement

The authors have declared no competing interest.

### Funding Statement

Non-funded work

